# Systematic disease-agnostic identification of therapeutically actionable targets using the genetics of human plasma proteins

**DOI:** 10.1101/2023.06.01.23290252

**Authors:** Mohd Anisul Karim, Bruno Ariano, Jeremy Schwartzentruber, Juan Maria Roldan-Romero, Edward Mountjoy, James Hayhurst, Annalisa Buniello, Elmutaz Shaikho Elhaj Mohammed, Miguel Carmona, Michael V Holmes, Chloe Robins, Praveen Surendran, Stephen Haddad, Robert A Scott, Andrew R. Leach, David Ochoa, Joseph Maranville, Ellen M. McDonagh, Ian Dunham, Maya Ghoussaini

## Abstract

Proteome-wide Mendelian randomization (MR) has emerged as a promising approach in uncovering novel therapeutic targets. However, genetic colocalization analysis has revealed that a third of MR associations lacked a shared causal signal between the protein and disease outcome, raising questions about the effectiveness of this approach. The impact of proteome-wide MR, stratified by cis-trans status, in the presence or absence of genetic colocalization, on therapeutic target identification remains largely unknown.

In this study, we conducted genome-wide MR and cis/trans-genetic colocalization analyses using proteomic and complex trait genome-wide association studies. Using two different gold-standard datasets, we found that the enrichment of target-disease pairs supported by MR increased with more p-value stringent thresholds MR p-value, with the evidence of enrichment limited to colocalizing cis-MR associations.

Using a phenome-wide proteogenetic colocalization approach, we identified 235 unique targets associated with 168 binary traits at high confidence (at colocalization posterior probability of shared signal > 0.8 and 5% FDR-corrected MR p-value). The majority of the target-trait pairs did not overlap with existing drug targets, highlighting opportunities to investigate novel therapeutic hypotheses. 42% of these non-overlapping target-trait pairs were supported by GWAS, interacting protein partners, animal models, and Mendelian disease evidence. These high confidence target-trait pairs assisted with causal gene identification and helped uncover translationally informative novel biology, especially from trans-colocalizing signals, such as the association of lower intestinal alkaline phosphatase with a higher risk of inflammatory bowel disease in *FUT2* non-secretors.

Beyond target identification, we used MR of colocalizing signals to infer therapeutic directions and flag potential safety concerns. For example, we found that most genetically predicted therapeutic targets for inflammatory bowel disease could potentially worsen allergic disease phenotypes, except for *TNFRSF6B* where we observed directionally consistent associations for both phenotypes.

Our results are publicly available to download or browse in a web application enabling others to use proteogenomic evidence to appraise therapeutic targets.

## Introduction

Many candidate drugs are supported by *in vitro* evidence and animal models yet fail during human clinical trials, motivating the pursuit of alternative sources of evidence to appraise therapeutic targets^1^. A major source of evidence is human genetic data linked to disease traits, which can identify targets more likely to be successfully modulated in clinical trials^2, 3^. However, disease-associated genetic signals often span multiple genes, and do not immediately indicate the desirable direction of therapeutic modulation^4, 5^. Selection of target genes can be improved by machine learning algorithms trained on curated gold standard target-trait datasets, but such datasets are biased towards the nearest gene and to known mechanisms such as protein-altering variants^6–8^.

Proteins are the most common therapeutic targets. The emergence of genetic data on plasma proteins has provided an alternative route to assign causal genes to genetic signals with high confidence, to help uncover novel disease biology and to inform the direction of therapeutic and adverse effects^9, 10^. For example, proteogenomic data were crucial to determining the beneficial direction of therapeutic effect of *OAS1* in COVID-19 disease^11^. Proteogenomic evidence also nominated circulating CD209 as one of the causal proteins that was associated with the COVID-19 linked *ABO*-locus^12^; subsequent studies provided experimental evidence of interaction of CD209 with the spike protein of the SARS-CoV-2 virus^13^.

Whereas there are numerous efforts to use similar proteogenomic evidence to identify actionable therapeutic targets, studies to systematically characterize proteogenomic evidence, especially using trans-acting genetic instruments, are limited^14–16^. In these previous studies, genetically predicted protein abundance was linked to disease-relevant traits by approaches such as Mendelian randomization (MR) and genetic colocalization (‘coloc’). The MR approach is based on the concept that, at the population-level, alleles are randomized at conception, enabling comparisons of groups in a population that differ only in the distribution of a genetically associated exposure trait analogous to the randomization of study participants to intervention and control arms. This enables investigators to create genetic instruments to represent effects of, for example, pharmacological modulation of the protein, and has been shown to recapitulate results of clinical trials^17^. However, Zheng et al^14^ showed that a third of proteogenetic MR associations are not supported by genetic colocalization, i.e. at a given region, the MR association of the protein trait with the disease trait was more likely to be driven by independent genetic signals than a shared signal and the MR association was likely genetically confounded by linkage disequilibrium. With a small test set of approved drug targets, Zheng et al also showed that target-trait pairs supported by combined MR-coloc evidence (P < 3.5 x 10^-7^ and H4 > 0.8) were slightly more enriched in approved drug targets than pairs not supported by combined MR-coloc (fisher’s exact test p = 0.05)^14^.

## Results

### Study overview

We used seven publicly available human blood plasma proteogenomic datasets^9, 18–23^ and genome-wide association studies (GWAS) of 3,766 complex traits from Open Targets that had at least one genome-wide significant locus (**Supplementary table 1 and 2**). We performed two-sample MR using genome-wide significant (5 x 10^-8^) instruments from proteomic GWAS (‘pan-MR’) and all available phenotypic GWAS, as well as cis/trans genetic colocalization to connect targets with traits (**Figure 1**). For MR analysis, although the presence of a genome-wide significant locus in the outcome data was a criteria for selection of outcomes, it is not necessarily the same locus that would be MR-analyzed; this would only be determined by the locus that is genome-wide significant in the proteomic GWAS. At a 5% false discovery rate (FDR; corresponding to MR p < 0.0005) across 2,276,452 target-trait pairs tested, there were 45,851 MR estimates representing 24,208 unique target-trait pairs. Of these, 21% were ‘cis-MR’ (based on instruments within 1 Mb of the transcription start site of the target protein), 58% were ‘trans-MR’ (instruments > 1 Mb from the target protein), and 21% had both cis- and trans-acting instruments (mixed MR). Cis-target-trait pairs from the Open Targets Genetics portal that underwent genetic colocalization following evidence of credible set overlap were merged with the cis-MR results. Almost half (49.3%) of cis-MR target-trait pairs had genetic colocalization results with only 32% of cis-MR target-trait having a posterior probability H4 > 0.8 (the typical threshold for evidence for genetic colocalization). Of the remaining cis-target-trait pairs with H4 < 0.8 with, the majority (98.8%) had higher H3 (evidence for independent signals) than H1 (genetic association with protein, but not with outcome) supporting reports from previous cis-MR-only studies of widespread genetic confounding^14^. However, since only significant loci from protein and outcome GWAS were analyzed for credible set overlap in our genetics portal to determine if a genetic colocalization test would be done, we cannot completely exclude the possibility that insufficient statistical power in the cis-MR results was responsible for the lack of genetic colocalization.

**Figure 1:**
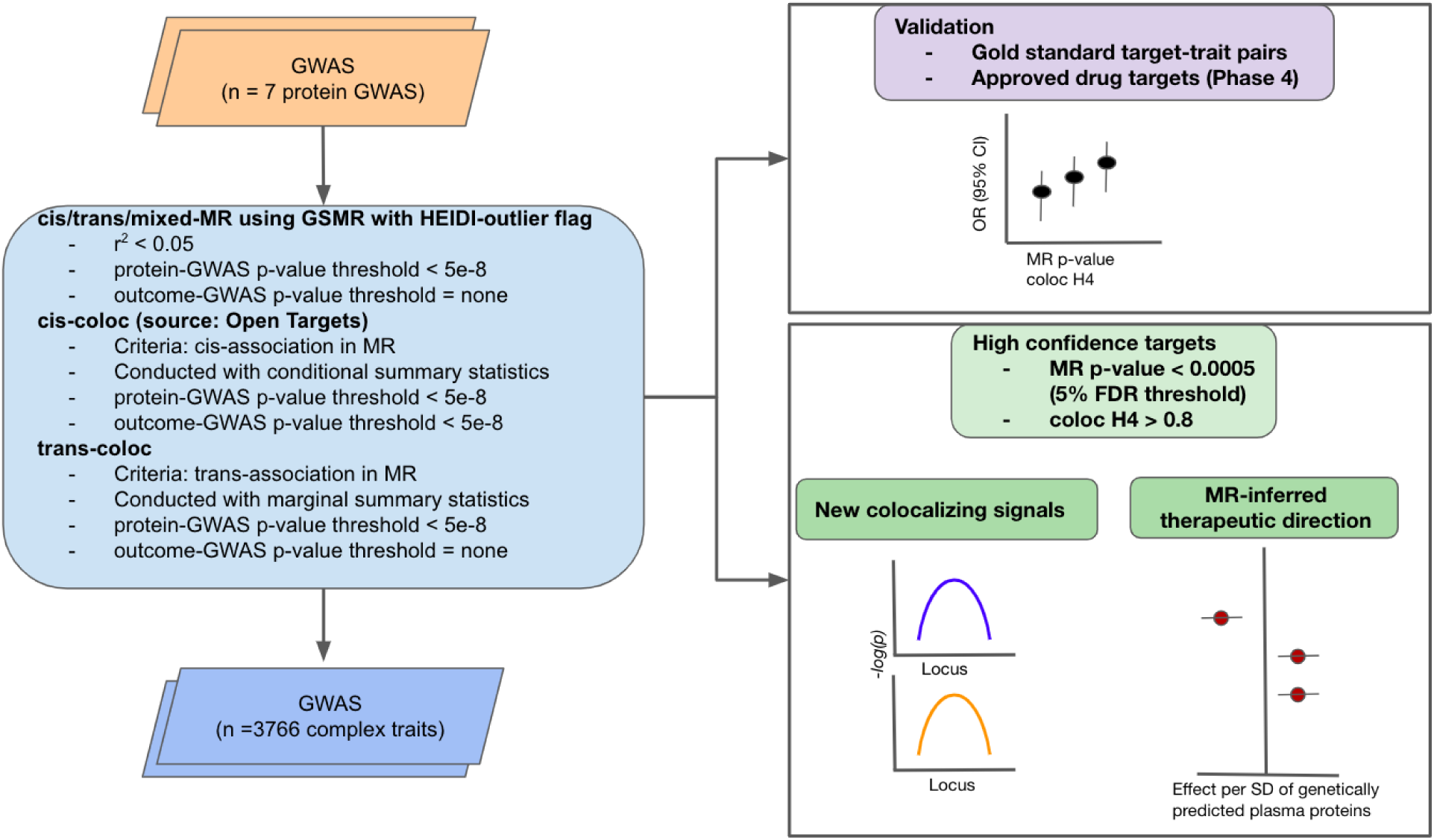
Study overview. The figure illustrates our systematic approach of using GWAS of proteins and complex traits. We used seven protein GWAS that were publicly available at the time of the analysis and 3766 complex trait outcomes selected on the basis of presence of a genome-wide association signal (as proxy for power). We performed genome-wide Mendelian randomization (MR), integrated the MR results with cis-colocalization results from the Open Targets Genetics portal and additionally performed trans-colocalization with trans-MR associations. We validated the results with gold standard datasets which informed our approach on curating a ‘high-confidence’ target-trait dataset. We generated examples of how this new dataset can be used, including showing potentially novel colocalizing signals and its use in inferring therapeutic directions. GWAS - Genome-wide association studies. MR - Mendelian Randomization. FDR - False discovery rate. coloc - genetic colocalization. cis-coloc/MR - if the gene near the associated/colocalizing loci encodes the protein in the GWAS used for MR/genetic colocalization. trans-coloc/MR - if the gene encoding the protein in the GWAS used for MR/genetic colocalization is not near the associated/colocalizing loci. Mixed-MR - when an MR association has both cis- and trans-acting genetic instruments.

For validation, we investigated evidence of enrichment in two different gold standard datasets of different pQTL-based trait associations (i.e. cis, trans, and mixed cis-trans) at different MR p-values with and without genetic colocalization evidence. First, we used an updated EFO-annotated clinical trial dataset recently incorporated in the Open Targets platform as the source of approved drug targets^24^. Second, we leveraged our ‘gold standard’ gene set (gene targets linked to traits with high confidence, of which 19% target-trait pairs overlap with target-trait pairs in the approved targets dataset) that was used to develop our locus-to-gene (L2G) classifier^6^.

### Evidence of enrichment is limited to cis-colocalizing target-trait pairs

We found progressively stronger enrichment of target-trait pairs in both the gold-standard gene set and approved drug targets with increasingly stringent MR p-values, and this was largely driven by cis-MR associations (**Figures 2A and 2B**, **Supplementary Tables 3, 4, 5 and 6**). To check whether this was because the trans- and mixed-MR associations were more statistically pleiotropic than cis-MR associations, we carried out MR-Egger and MR-Weighted Median analyses on all MR associations. We found the effect estimates from GSMR and MR-Egger/Weighted Median approaches to be highly correlated (r2 > 0.9) in a similar manner in cis-, trans, and mixed-MR associations suggesting that pleiotropic associations were unlikely to explain the higher enrichments with cis-MR associations compared to trans- or mixed-MR associations (**Supplementary Figure 1**). Moreover, when we examined cis-MR target-trait pairs by their genetic colocalization status (cis_coloc_H4 > 0.8), we observed limited variation in enrichment by progressively stringent MR p-value (**Figure 2C**, **Supplementary Tables 7 and 8**), emphasizing the important role of genetic colocalization for therapeutic target identification alongside MR which is better suited for assessment of directionality of therapeutic effect.

**Figure 2:**
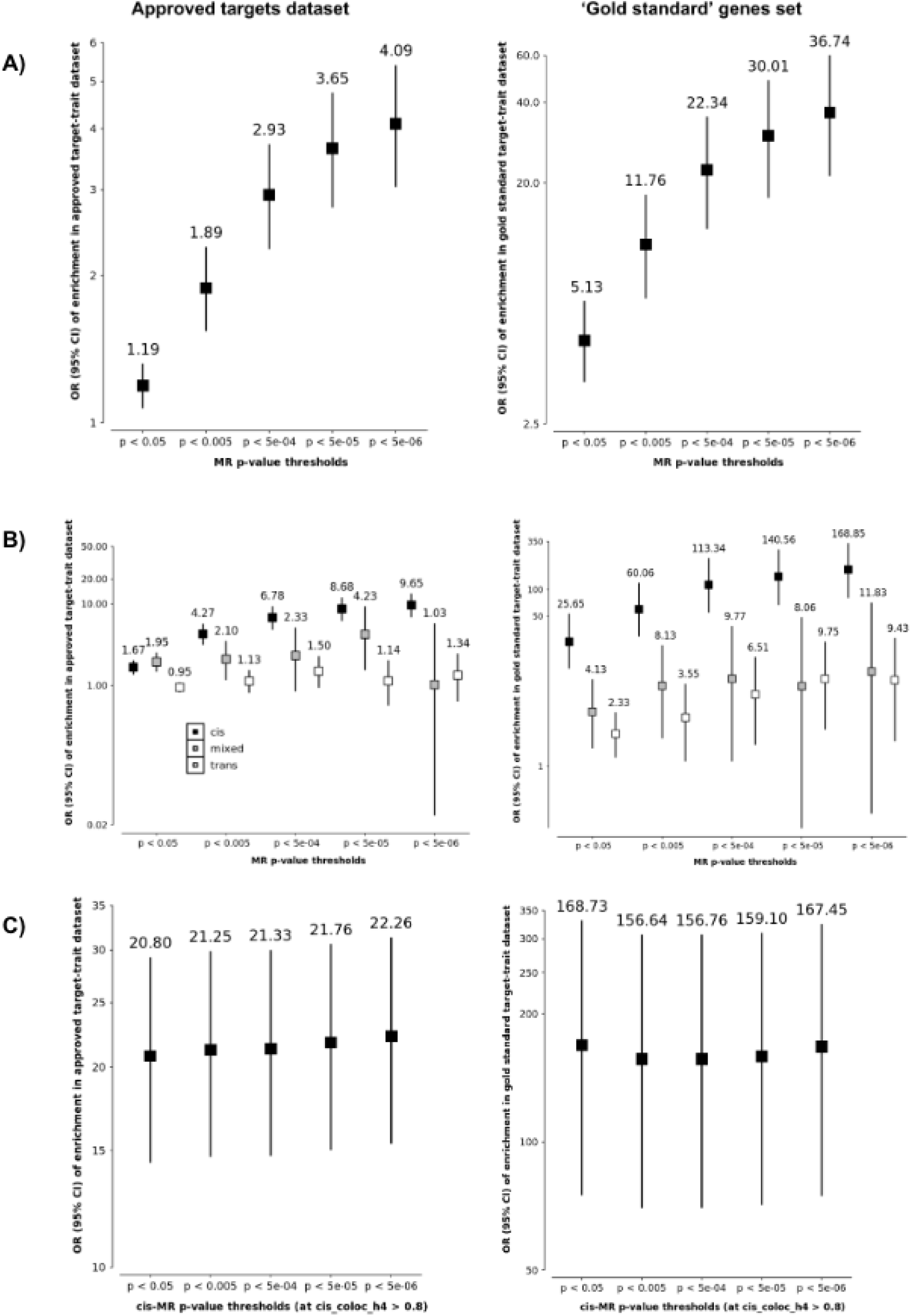
Enrichment of MR predicted target-traits in the approved drugs and Open Targets gold standard validation sets . The y-axis shows the odds ratio which estimates the odds of the associated target-trait pair being part of the gold-standard datasets compared to the odds of target-trait pairs below (colocalization/coloc H4) or above (MR p-value) the designated thresholds. A) Evidence of progressive enrichment at higher MR p-value thresholds, irrespective of genetic colocalization evidence, B) Breakdown of enrichment by cis-trans-mixed MR status, showing progressive enrichment driven by cis-MR predicted target-trait pairs, and C) Within cis-MR results, there is limited variation of enrichment when considering colocalizing target-trait pairs (H4 > 0.8) (cis-MR only results are tabulated in Supplementary tables 7 and 8)

### Phenome-wide proteogenetic colocalization and Mendelian randomization identifies new high confidence potential drug targets and uncovers novel biological pathways

The results from MR and genetic colocalization (MR-coloc) were used to curate a high confidence target-trait dataset (MR p-value < 0.0005 corresponding to 5% FDR and colocalization H4 ≥ 0.8) of 774 unique target-trait pairs, of which 285 pairs were from cis-coloc associations (of which half are linked to protein-altering variants), and 489 pairs were from trans-coloc associations (these are proteins with trans or mixed MR associations with a trait where there is evidence of genetic colocalization for at least one of the trans-pQTL signals and the trait) (**Figure 3**, **Supplementary Table 9**). Of the 774 target-trait pairs, 202 pairs show evidence of association in more than one outcome dataset, 69 pairs in more than one proteomic dataset, and 32 pairs replicate in both proteomic and outcome datasets.

**Figure 3:**
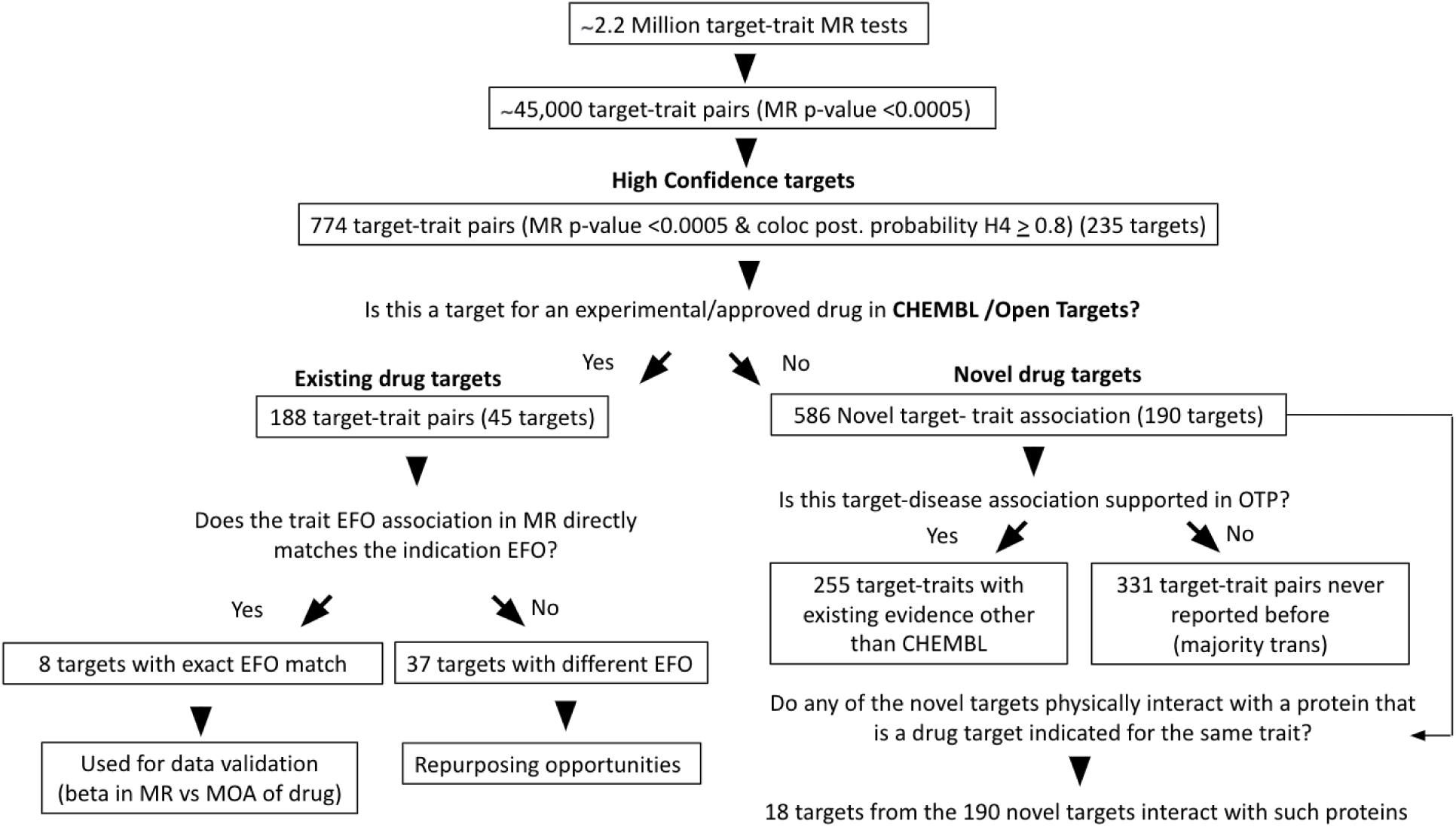
Workflow illustrating the logic of the different research questions we asked and classification schemes applied for existing and novel target-trait pairs. For the 255 target-traits with existing evidence other than ChEMBL, we examined for evidence from different sources used by the Open Targets Platform (e.g. GWAS evidence, animal models, RNA expression atlas, Mendelian disease evidence, and text-mined literature). To check for protein-protein interaction, we used the IntAct database with a stringent threshold (molecular interaction score > 0.42). MR - Mendelian randomization; coloc - genetic colocalization; EFO - Experimental Factor Ontology; MOA - Mechanism of Action.

Of the 774 target-trait pairs, 188 pairs (68 pairs being cis) involved 45 known drug targets while the remainder (586 pairs involving 190 targets) were classified as ‘novel’ targets. 255 of both cis or trans pairs out of the 586 ‘novel’ target-trait pairs were also supported by other sources of evidence from the Open Targets platform which includes the machine-learning ‘locus-to-gene’ (L2G) method^6^, animal models, RNA expression atlas, Mendelian disease evidence, and text-mined literature. Of the remaining 331 target-trait pairs that were not supported by other evidence sources, the majority of these were trans target-trait pairs.

Using the IntAct database of physical protein-protein interaction, we identified 18 targets from the 190 novel targets where the protein they interacted (molecular interaction score > 0.42) with was a drug target indicated for the colocalizing MR-supported trait. These colocalizing target-drug target pairs included several receptor-ligand pairs (e.g. IL1RL1-IL33 for asthma, IL23R-1L23 for IBD) and enzyme-substrate pairs (REN-AGT for hypertension). Some interactions suggested biological pathways that could mediate the genetically predicted therapeutic effect. For example, the association of pancreatic lipase related protein 1 encoded by the pancreas-specific *PNLIPRP1* gene with type 2 diabetes may be related to mitochondrial complex I (*NDUFAF1*) inhibition given that *NDUFAF1* (that interacts with *PNLIPRP1*) is one of the targets of metformin^25^ (**Supplementary Figure 2**) Another example is the interaction of *CD209* - a pathogen-recognition receptor expressed on dendritic immune cells and associated with COVID-19 disease, with *CASP6* (caspase 6) - a cysteine protease that contributes to antiviral host defense by acting as a central mediator of pyroptosis, apoptosis, and necroptosis (PANoptosis) of virus-infected cells^26^. Caspase 6 is one of the targets of the pan-caspase inhibitor emricasan that was recently in Phase 1 trials for COVID-19 but now terminated due to problems in recruitment (NCT04803227). Nevertheless, the *CD209-CASP6* link implicates the apoptosis pathway as therapeutically relevant for COVID-19, and supports our previous observation of genetic colocalization of soluble Fas with COVID-19 disease^13^; the binding of Fas with Fas ligand on cells activates the caspase cascade that initiates apoptosis. These examples suggest that by harnessing protein-protein interactions, it is possible to not only uncover new biological insights but also effectively broaden the range of potential druggable targets.

### Colocalizing target-trait pairs recapitulates mechanism of action of drug targets

To determine whether the high confidence target-trait dataset can reliably inform therapeutic direction, we examined whether the direction of the genetically predicted therapeutic effect of the proteins matches the mechanism of action of drugs at different phases of drug development (**Supplementary Table 9**). Of the 188 pairs representing 45 drug targets, there were 8 targets where the associated trait EFO code directly matched the indication EFO (**Figure 3**, **Supplementary Table 9**). Six out of the 8 targets were supported by cis-coloc associations (*IL12B, IL2RA, PCSK9, TNFSF11, F2, APOB*) and only two targets (*IL2RB, INSR*) were supported by trans-coloc (*SH2B3, ABO*). For cis-targets, we found that the genetically predicted therapeutic direction was consistent with the mechanism of action for 5 out of the 6 targets, except for the association of apolipoprotein B with hypercholesterolemia. For the latter, genetically predicted higher apolipoprotein B was associated with a lower risk of hypercholesterolemia, inconsistent with the biological role of apolipoprotein B and opposite to the mechanism of action of Mipomersen (an mRNA antisense inhibitor). However, the mismatched apolipoprotein B genetic association was only present in aptamer-based proteomic datasets and not observed in other lipoprotein GWAS where apolipoprotein B was measured using, for example, immunoturbidimetric methods as in the UK Biobank^27^. Furthermore, the *APOB* signal was also a cis-eQTL in adipose tissues (https://genetics.opentargets.org/variant/2_21036690_T_C) and when examining the direction of the cis-eQTL, it was also opposite to what was predicted by aptamer-based cis-pQTL. The directional mismatch may be due to the cis-pQTL signal (rs1469513) being in linkage disequilibrium (LD r^2^ = 0.6) with the missense variant (rs679899) in the *APOB* gene (i.e. an aptamer-binding artifact). This example emphasizes that when cis-pQTLs are protein-altering variants (PAVs) or in high LD with PAVs, the genetically predicted therapeutic direction should be interpreted in the context of other sources of information like known biology, non-aptamer-based protein quantification methods, and cis-eQTLs in disease-relevant tissues. In the absence of known biology of the trans-associations, there is a high level of uncertainty inferring whether there is a match between the genetically predicted therapeutic direction of the protein and the mechanism of action of the drug for a particular trans-target-trait association. Nevertheless, we found that 5 out of 6 *IL2RB* linked target-trait associations were consistent with the expected mechanism of action of aldesleukin, i.e. genetically predicted lower soluble interleukin-2 receptor beta that is associated with the therapeutic effect represents the agonistic action of interleukin-2 indicated for the same traits (autoimmune disease and coronary heart disease). However, there was inconsistency of the associations of soluble interleukin-2 receptor beta with breast cancer and insulin receptor with hypercholesterolaemia, (for detailed notes on how trans/cis-target therapeutic directions were manually assessed, see column ‘notes_if_predicted_matches_indicated_target_trait_pair’ in **Supplementary Table 9**).

### Trans-colocalizing target-trait pairs are informative of actionable therapeutic biology

Separately, although there was limited evidence of enrichment of trans-associations in either of the datasets, when we examined the individual trans-associations, we uncovered several biologically plausible and informative target-trait associations.

For example, there was evidence of genetic colocalization of the trans-association between CCL21 protein levels (encoded by *CCL21* on chromosome 9) and total cholesterol level, with the signal located near the *ACKR4* gene region on chromosome 3 (**Figure 4**). The ACKR4-CCL21 trans-association is biologically plausible given that the atypical chemokine receptor ACKR4 acts as a decoy receptor that sequesters and degrades chemokines like CCL21 controlling immune cell chemotaxis^28^ and implicates the ACKR4-CCL21 chemotactic inflammation pathway in influencing cholesterol levels.

**Figure 4:**
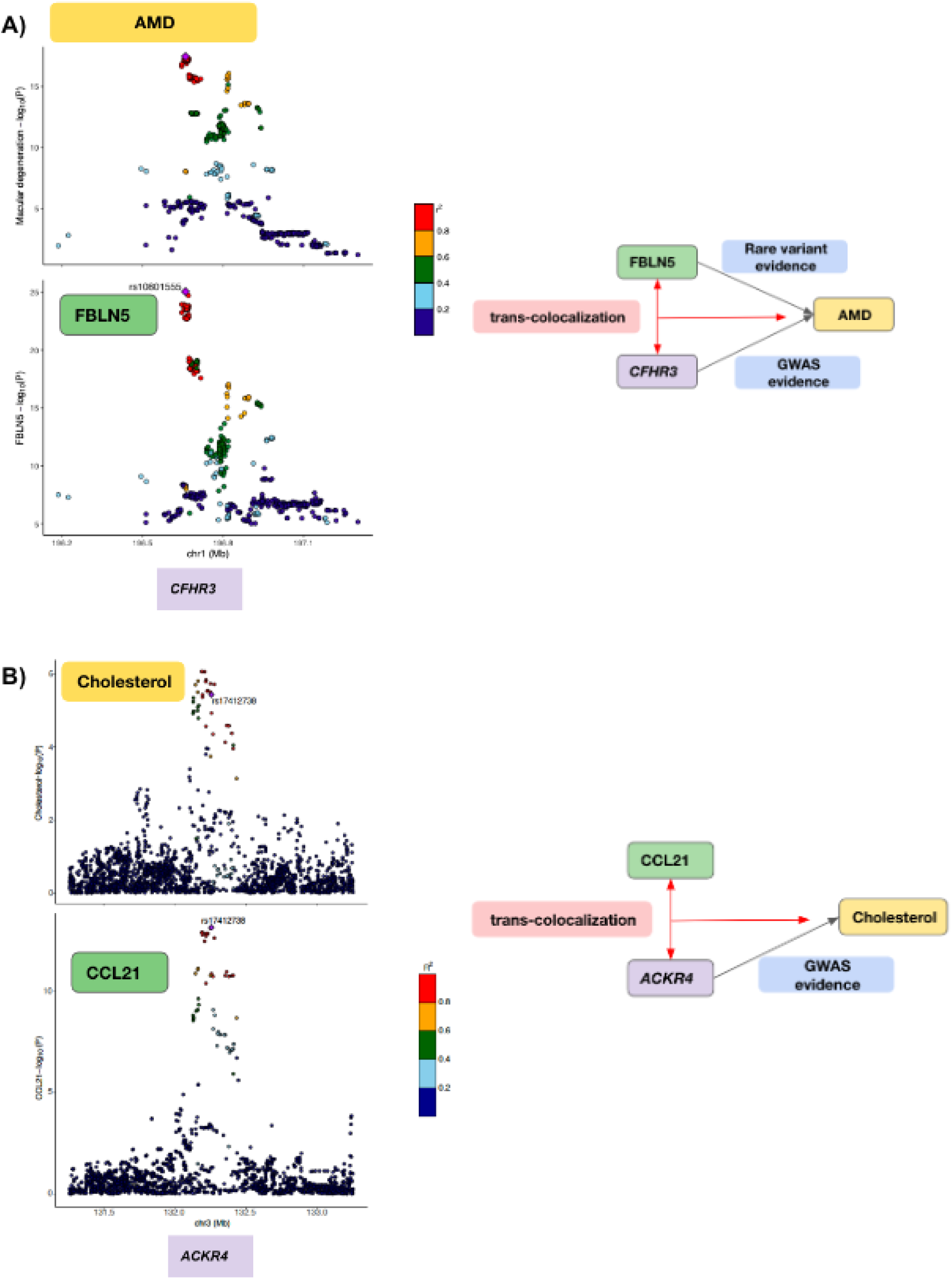
Genetic colocalization of trans-MR association between A) FBLN5 protein and acute macular degeneration (AMD) near *CFHR3* gene B) CCL21 protein and total cholesterol near the *ACKR4* gene.

Additionally, there were two novel colocalizing trans-associations. First, the colocalizing signal (rs10801555) near one of the complement factor H (CFH) genes (*CFHR3*, chromosome 1) supporting the trans-association between fibulin-5 (*FBNL5*, chromosome 14), a member of the fibulin family of extracellular matrix proteins and age-related macular degeneration (AMD).

While the CFH association with AMD is established through GWAS, the connection with fibulin-5 was only supported by rare variant evidence^29, 30^. However, the interaction of the complement system of proteins with fibulin (specifically fibulin-3) has been demonstrated in in vivo cellular and animal models^31, 32^. The present *CFH* locus-driven colocalizing association between FBLN5 and AMD raises the question of whether soluble fibulin-5 should be considered a biomarker of drugs modulating the complement system (e.g. recombinant human CFH therapy^33^) or a drug target in its own right.

Second, a colocalizing signal (rs516246) near the fucosyltransferase 2 gene (*FUT2*, chromosome 19) is in near-complete linkage disequilibrium (r2 = 0.99) with a stop-gain mutation in *FUT2*, and its trans-MR association genetically associates higher intestinal alkaline phosphatase (*ALPI*, chromosome 2) with a lower risk of inflammatory bowel disease (IBD) (**Supplementary Figure 3**). A number of studies (including GWAS) have shown that *FUT2* non-secretor status is a risk factor for IBD^34^. However, there is, to date, no GWAS evidence supporting the direct association of ALPI with IBD. Despite this, biallelic mutations in *ALPI* are a Mendelian cause of IBD^35^, and *ALPI* gene expression is lower in IBD patients compared to healthy controls in the RNA expression atlas (https://platform.opentargets.org/evidence/ENSG00000163295/EFO_0003767). The inferred direction of therapeutic effect where higher levels of ALPI associate with lower IBD risk, also support the results of a small open-label clinical trial that demonstrated benefit of exogenous alkaline phosphatase on ulcerative colitis

### Colocalizing target-trait pairs nominate likely causal genes at unresolved loci

We used the curated high confidence therapeutic targets dataset to resolve likely causal genes for binary traits where the identity of the causal gene remained unclear using our L2G prediction (i.e. either the L2G score was absent or less than 0.5) (**Supplementary Table 10**). The 774 unique target-trait pairs in the high confidence dataset represented 235 unique targets and 168 unique EFO traits, from which there were 207 targets for 124 EFO traits where the L2G score was absent or less than 0.5. In the latter, about 86 targets associated with 82 EFO traits were also supported from other sources of evidence like text-mined literature as being associated with the trait. Such examples include known associations of lower agouti-signaling protein (*ASIP*) with skin cancer^36^ or higher chitotriosidase (*CHIT1*) being a marker of asthma protection at the *ADORA1* locus^37^. Some associations were supported only by animal studies. For example, a trans-colocalizing signal at rs55993634 (*CTRB2*, chromosome 16) that associated genetically predicted lower pancreatic lipase related protein 1 (*PNLIPRP1*, chromosome 10) with lower risk of type 2 diabetes but higher risk of type 1 diabetes - both *CTRB2* and *PNLIPRP1* genes are expressed exclusively in the pancreas and the association of PNLIPRP1 with type 2 diabetes is supported by knockout studies in animal models^38, 39^. Some target-trait pairs were supported with both animal model and preliminary clinical trial evidence such as the association of lower plasma myeloperoxidase (*MPO*) protein with lower risk of cardiovascular disease^40, 41^. Many of the low L2G-scoring target-trait associations revealed by MR-coloc were unique to specific population groups but biologically plausible. For example, a signal near the gene cluster *MTHFR-CLCN6-NPPA-NPPB* (nearest gene: *CLCN6*) associated with higher B-type natriuretic peptide (*NPPB*) colocalized with a lower risk of pregnancy-induced hypertension-related phenotypes in Finns^42^. Some target-trait associations from MR-coloc were novel, i.e they were not supported by other evidence sources from the Open Targets platform, e.g. *IGHG1* (immunoglobulin heavy constant gamma 1) and hypothyroidism, and, therefore, provide new hypotheses for future research.

### Mendelian randomization of colocalizing target-trait pairs suggested *TNFRSF6B* as a therapeutic target for IBD and allergic disease phenotypes

We demonstrate that, for targets with evidence of efficacy for a disease, our approach can be used to reliably infer potential adverse events and identify alternative targets. As an example, we show that several genomically-informed therapeutic targets for inflammatory bowel disease were also associated with a higher risk of allergic disease phenotypes. We propose an alternative IBD/allergic disease phenotype target *TNFRSF6B* that was genetically predicted to have the same direction of effect for both IBD and allergic diseases (**Figure 5**, **Supplementary Figure 4**).

**Figure 5:**
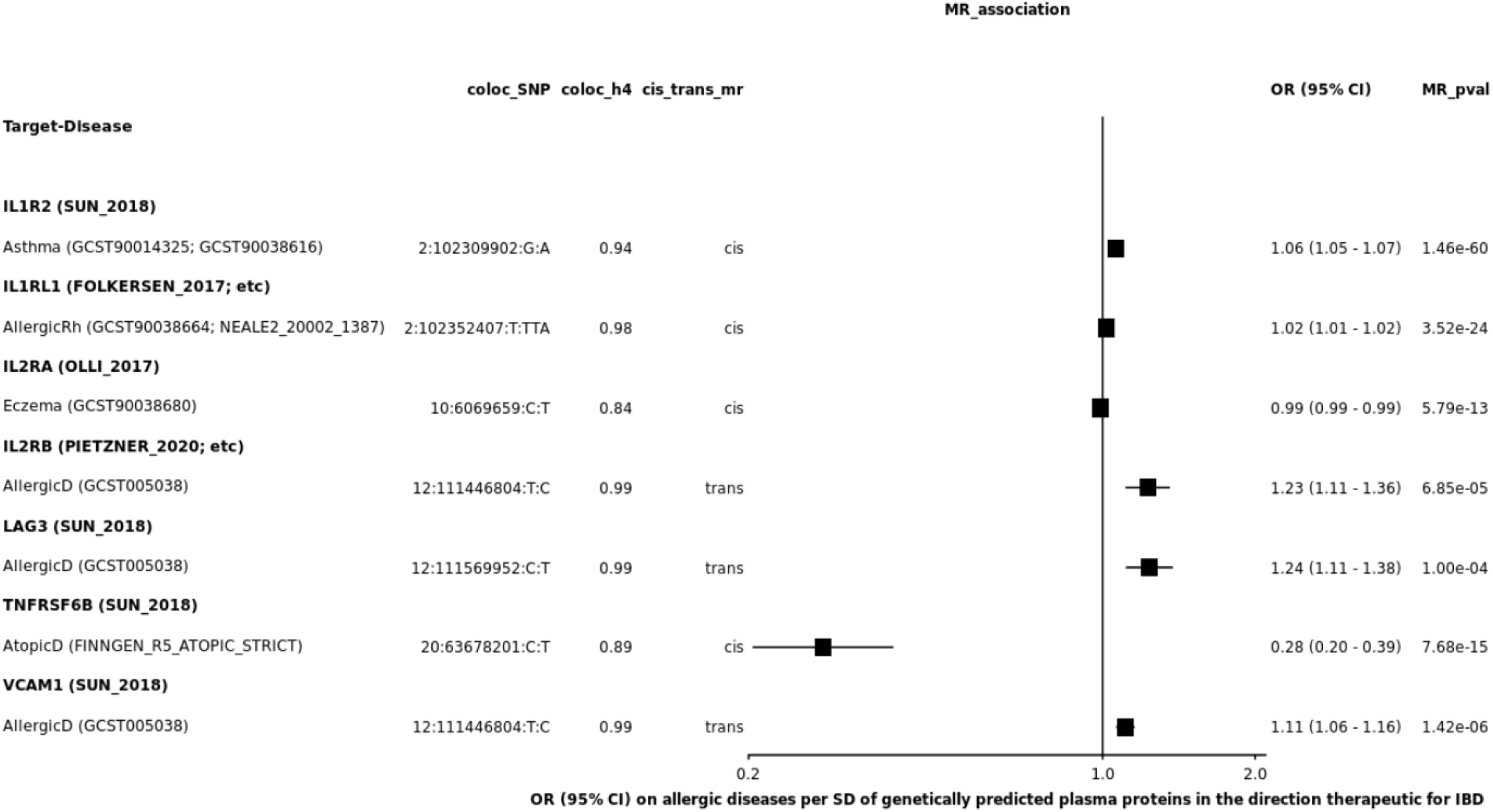
Association of colocalizing genetically predicted soluble proteins with allergic disease phenotypes (associated study IDs for reference) oriented in the direction that is therapeutic for inflammatory bowel disease (IBD). For cis-MRs, we ensured the cis-acting coloc_SNPs were not protein-altering or in linkage disequilibrium (r2 >= 0.5) with protein-altering variants for their respective protein targets. The full figure showing directions for both allergic disease phenotypes and IBD is provided in the Supplement (Supplementary Figure 4). **AllergicRh - Allergic rhinitis; AllergicD - Allergic disease; AtopicD - atopic dermatitis coloc_SNP - SNP representing the colocalising signal; coloc_H4 - posterior probability for a shared causal signal; cis_trans_mr - whether the MR association is from cis-acting or trans-acting SNPs; OR (95% CI) - Odds ratio and 95% confidence interval; MR_pval - p-value for the MR association.**

## Discussion

We investigated the relative contributions of proteomic MR and genetic colocalization, including their cis-trans status, to enrichment of target-trait pairs in gold-standard datasets. We provide empirical evidence demonstrating the importance of genetic colocalization with MR association to identify therapeutic targets. Although enrichment was limited to cis-target-trait pairs, when informed by biological mechanism and additional lines of evidence, the trans-colocalizing signals were biologically informative and we showcased three examples to illustrate their potential translational relevance. Two of the trans-colocalizing signals (nearest trans-gene: *CFHR3*, protein: fibulin-5, disease: AMD and *FUT2*, intestinal alkaline phosphatase, IBD) had no direct GWAS evidence and were not identified in previous proteomic MR-coloc studies. Overall, our approach yielded 774 high confidence target-trait pairs at thresholds of coloc_h4 > 0.8 and an MR p-value < 0.0005 (equivalent to 5% FDR), with 24% of these matching known drug targets. Of the novel targets, 42% were supported by either GWAS, animal models, or Mendelian disease evidence. Eighteen of the novel targets interacted with known drug targets that have trait-matching indications, providing opportunities to formulate novel therapeutic hypotheses to explain their genetically predicted therapeutic effect.

A key strength of our study is the systematic approach we used to include proteomic and outcome studies agnostic to any particular therapeutic area, which contrasts with most previous work that has used focused proteomic and outcome GWAS^14–16^. Specifically, we expand on previous work in five major ways. First, by separately analyzing enrichments of MR-selected and coloc-selected target-trait pairs including by their cis-trans status, we were able to assess the relative contributions of proteomic MR and genetic colocalization to appraising therapeutic targets. Second, the use of larger gold-standard datasets enabled us to demonstrate stronger enrichment at progressively more stringent MR and colocalization thresholds. Third, we triangulated evidence supporting colocalizing target-trait pairs from other non-GWAS sources, identified colocalizing target-trait pairs that replicate in different protein or outcome datasets, and used protein-protein interactions to identify protein partners that are drug targets for the indication matching the colocalizing trait, all of which provides additional confidence to our results. Fourth, we selected a larger number of outcome datasets agnostic to any specific therapeutic areas which enabled us to build a bigger list of colocalizing high confidence target-trait pairs than, for example Zheng et al (489 vs 270, using their stringent MR p-value cut-off 3.5 x 10^-7^) with only 13% target overlap with our dataset (**Supplementary Table 13**). Finally, we used expert assigned EFO codes to match colocalizing target-trait pairs with drug target-indication pairs as opposed to using a similarity matrix derived from MeSH headings in Zheng et al which can be sensitive to different cut-offs^2^. However, we should also note a limitation from our end that the use of EFO to count unique EFO IDs will likely contain duplicate traits and miss related quantitative traits.

Our study has several limitations. Our associations are limited to publicly available GWAS summary statistics available via Open Targets which is limited both in terms of larger proteomic studies and disease outcomes, in particular cancer phenotypes^43^. The summary statistics used in our study represent associations of common variants in populations where the majority are of European descent. Increasing the representation of ethnically diverse populations in future genetic studies is likely to capture additional novel signals from genetic variants that are otherwise rare in the European populations and fuel therapeutic target discovery campaigns. Additionally, the Olink/Somalogic platforms used by the respective studies cover circulating plasma proteins only which may not necessarily be the disease-relevant tissue, capture less than a fifth of the human proteome and are also unable to distinguish free from bound protein, limiting the interpretation of mechanistic insights and assessment of pleiotropic associations. Further, as noted by previous studies, affinity-based platforms for protein measurement rely on the shape of the canonical protein to estimate protein abundance and can miss genetic effects specific to a particular isoform of the protein^44^, misrepresent direction of effects when genetic instruments for MR are PAVs or linked to PAVs, or lead to false negative associations. For the latter, although we have performed MR using these instruments, we have appropriately annotated when these associations are driven by PAVs so that support for the directional information can be pursued from other sources of evidence as we have highlighted for the association between apolipoprotein B and hypercholesterolemia. The lack of enrichment for trans-pQTL based associations in our study may be due to insufficient power to detect an association as the generally lower effect estimates of trans-pQTLs relative to cis-pQTLs (**Supplementary Figure 5**) means we would require larger sample sizes to detect trans-pQTLs for us to then use them as instruments for MR analysis. Additionally, for computational capacity reasons, we used marginal summary statistics to perform trans-colocalization versus using conditional summary statistics as was done for cis-colocalization and this likely contributed to higher rates of false negative trans-colocalization results. Nevertheless, our work highlights the potential of trans-pQTL based associations to inform therapeutically actionable biology as exemplified by *FUT2-ALPI* associations on IBD.

We expect our systematic framework to be of value for upcoming larger proteogenomics projects, for example the recent industry-wide effort that assayed ∼1500 proteins on ∼55,000 UK Biobank (UKBB) study participants^45^, that can more reliably assess the translational value of both cis-pQTL and trans-pQTL based associations, with the ultimate aim of generating novel therapeutic hypotheses and improving the odds of clinical trial success.

### Online Methods

#### Genetic associations of proteomic data

We used seven publicly accessible proteogenomic datasets^9, 18–23^ for the pan-/cis-MR (cis = ±1Mbp from transcription start site) analyses and for performing genetic colocalization tests (described below). The pan-/cis-MR effects were expressed per standard deviation (SD) higher genetically predicted plasma protein concentrations. The genotyping protocols and QC of these proteomic studies have been described previously in the respective studies’ publications. Annotation of protein-altering variants or variants in linkage disequilibrium (LD) at an r2 = 0.5 was done using the TOP-LD tool^46^.

#### Genetic associations of complex trait data

We selected complex trait GWAS as outcomes if they had at least one genome-wide significant (p ≤ 5 x 10^-8^) locus, ensuring a systematic disease-agnostic trait selection strategy. This resulted in 3,766 traits, including traits from FinnGen (release 5), UK Biobank, and GWAS Catalog. A list of these studies are provided in **Supplementary Table 1**.

#### Harmonization of protein and outcome summary statistics

We used genomic coordinates based on the GRCh38 genome assembly; where required, we lifted over genomic coordinates from the GRCh37 assembly to GRCh38. We also ensured that the effect allele in a GWAS locus is the alternative allele in the forward strand of the reference genome and used a strand consensus approach to infer strand for palindromic variants (variants with A/T or G/C alleles or variants with the same pair of letters on the forward strand as on the reverse strand). Details of the harmonization workflow and the strand consensus approach are described in our previous publications^6, 13^ and documented with code in our GitHub pages (https://github.com/EBISPOT/gwas-sumstats-harmoniser; https://github.com/opentargets/genetics-sumstat-harmoniser).

#### Mendelian randomization

To construct genetic instruments for MR analysis, we used genome-wide (‘pan-MR’) significant near-independent (r-squared < 0.05) genetic instruments with no evidence of statistical heterogeneity and accounted for residual linkage disequilibrium (LD). The process of genetic instrument selection and MR analysis was automated using the generalized summary data-based Mendelian randomization (GSMR^47^) approach with the heterogeneity-independent instrument (HEIDI)-outlier flag turned on). The GSMR software, using the HEIDI-outlier method, removes potentially pleiotropic instruments and accounts for the residual correlation between instruments (important as we are using near-independent genetic instruments). To select near-independent genetic instruments and account for linkage disequilibrium (LD) in the MR analyses, we used genotype data from 10,000 randomly sampled UK Biobank participants to create a reference LD matrix. Additionally, for single SNPs representing cis-colocalising signals from our genetic colocalization pipeline (see below), we used the beta and standard error of the target-trait pairs to derive the Wald ratio which represented a single-instrument MR. We used a 5% false discovery rate using the Benjamini-Hochberg method as the MR p-value threshold to select high confidence therapeutic targets.

#### Genetic colocalization analysis

The full analysis methods of genetic colocalization of cis-pQTLs have been described in our previous publications^6, 8^. In brief, the full GWAS summary statistics of complex traits and cis-genetic regions of plasma proteins was used to identify conditionally-independent signals with genome-wide significance (p < 5e-8) using GCTA-COJO^48^; pairwise genetic colocalization was carried out using the *coloc* R package^49^ on the conditionally-independent summary statistics when at least one variant overlapped between credible sets for the two traits. For trans-MR associations, we used the full marginal summary statistics of each locus (not conditionally independent). The default priors in coloc were used, that is, the prior of a SNP (single nucleotide polymorphism)-trait association is 1 × 10^-4^, and the prior of a SNP associating with both traits is 1 × 10^-5^. For each target-trait pair, a posterior probability for shared causal genetic signal (H4) threshold of more than 0.8 was used to identify shared causal genetic variants and was the criteria to select high confidence therapeutic targets. The region corresponding to the MHC (put exactly what region this covered here, e.g. chr6:30-32 Mb) was excluded from this analysis.

#### Enrichment analyses

Ensembl IDs for genes and experimental factor ontology (EFO) IDs for traits were used to merge data from the approved drugs dataset (https://platform.opentargets.org/downloads, v22.11) and the GWAS gold standards (https://github.com/opentargets/genetics-gold-standards) with the target-trait dataset from MR and genetic colocalization analysis. For comparisons of MR and genetic colocalization with approved drug targets at different MR and colocalization thresholds, the 2 x 2 tables compared target-trait pairs that were phase 4 approved drug targets with any overlapping target-trait pair irrespective of their drug target status (**Supplementary Table 11**).

For comparisons with the larger gold standard positive target-trait pairs, the 2 x 2 tables compared target-trait pairs that were gold standard positive target-trait pairs with any overlapping target-trait pair irrespective of their gold standard positive status (**Supplementary Table 12)**. The cis-colocalization only analysis was limited to cis-colocalizing target-trait pairs irrespective of MR significance. The cis-MR only analysis was limited to cis-MR associations without evidence of genetic colocalization (H4 < 0.8). The Parent EFO IDs were used to map the traits in the case of approved drugs dataset and trait-specific EFO IDs were to map traits in the gold-standard positive dataset. Fisher’s exact test was used to compute p-values.

#### Protein-protein interactions analyses

Direct EFO terms were used for drug indications. To search for drugs targeting proteins that physically interact (henceforth ‘partner’) with each target under study, IntAct database was queried using a stringent threshold (molecular interaction score > 0.42), which means robust support for physical interactions as reported by the database and used by others^3, 50^. Self-interactions were not considered further for this analysis. When the partner drug had the same indication as the trait associated with the target under study, the name of the partner, the name of the drug and phase of clinical trial for the trait were annotated. To know if the target-trait pairs were supported by OT platform evidence, we annotated each with the direct associations scores from the OT platform and collated the data sources supporting the associations^24^.

## Data availability

Summary data for both proteins and outcomes used for genetic analyses are publicly available from the GWAS catalog https://www.ebi.ac.uk/gwas/downloads/summary-statistics.

Full results can be downloaded here:

FTP: https://ftp.ebi.ac.uk/pub/databases/opentargets/publishing/mendelian_randomisation_results/

Google cloud bucket: https://console.cloud.google.com/storage/browser/open-targets-genetics-releases/Mendelian%20randomisation%20results;tab=objects?authuser=1&project=open-targets-genetics&prefix=&forceOnObjectsSortingFiltering=false

Filtered results (bxy_pval < 0.0005 or coloc_h4_h3 > 1) can be browsed here: https://mk31.shinyapps.io/mr_app2/

## Code availability

https://github.com/opentargets/mendelian-randomisation

## Funding statement

MAK, BA, JS, JH, AB, DO, MC, EMM, MG, ID were funded by Open Targets. This research was funded in part by a Wellcome Trust [Grant number 206194]. For the purpose of Open Access, the authors have applied a CC-BY public copyright license to any Author Accepted Manuscript version arising from this submission.

## Author contributions

MAK, MG, and ID conceived the study. JH and AB harmonised all datasets. MAK performed all MR and enrichment analyses. EM and JS performed the cis-colocalization analyses. BA performed the trans-genetic colocalization analysis. JMRR conducted the protein-protein interaction analyses. MC provided analytical support to carry out analyses in Google cloud virtual machines. All authors provided valuable feedback and critical comments that informed the design and analyses in the present study.

## Competing interests

MAK is now an employee of Variant Bio. JS is now an employee of Illumina. JM is an employee of Bristol-Myers Squibb. ESEM is an employee of Genmab. EM is an employee at Genomics PLC. MVH is an employee of 23andMe, CR, PS, SH and RAS are employees of GlaxoSmithKline.

## Ethics declaration

All institutions contributing cohorts to the proteomics and outcome studies received ethics approval from their respective research ethics review boards.

## Data Availability

All data produced are available online at: https://ftp.ebi.ac.uk/pub/databases/opentargets/publishing/mendelian_randomisation_results/

