## Supplementary Figures for "Systematic disease-agnostic identification of therapeutically actionable targets using the genetics of human plasma proteins"

**Supplementary Figure 1: Effect estimate correlation plots between GSMR beta and MR-Egger/Weighted Median beta.**

MR-Egger

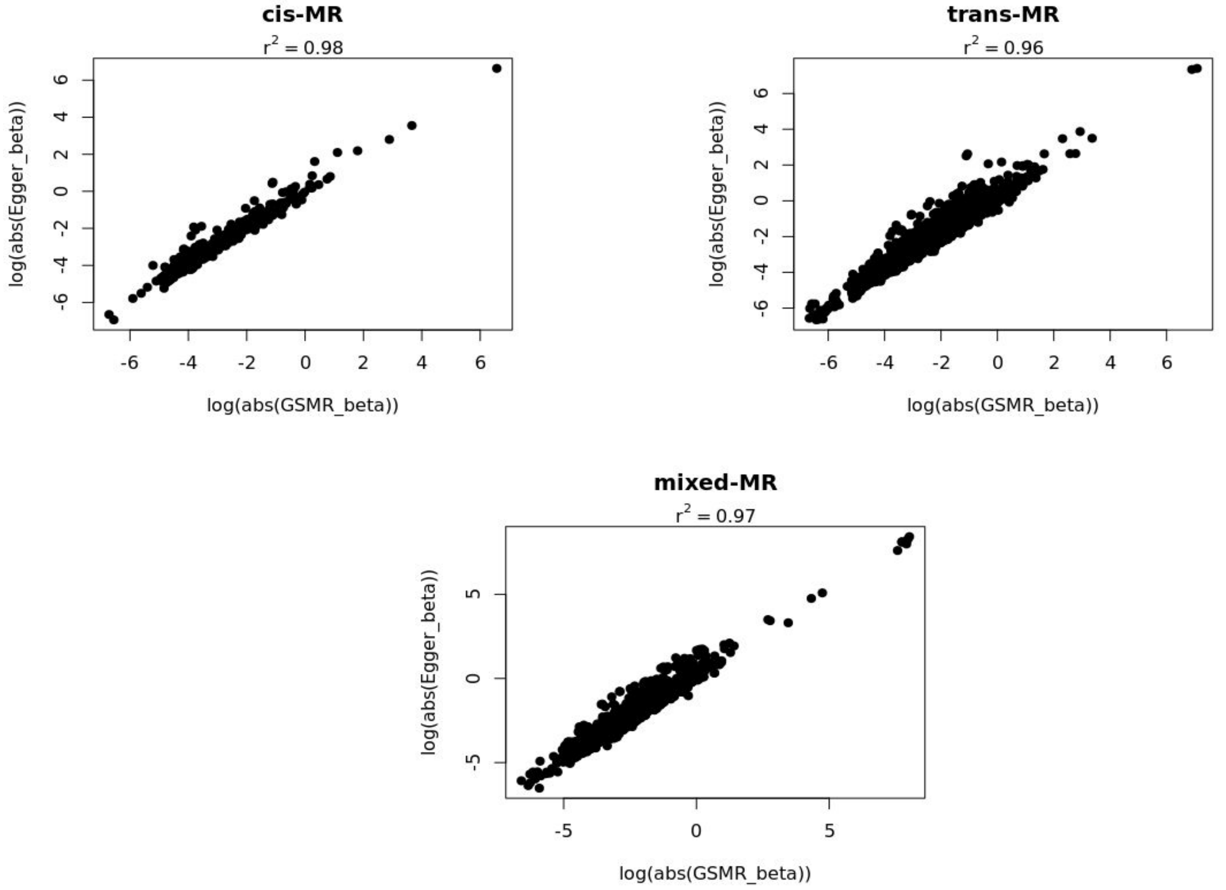


Weighted Median

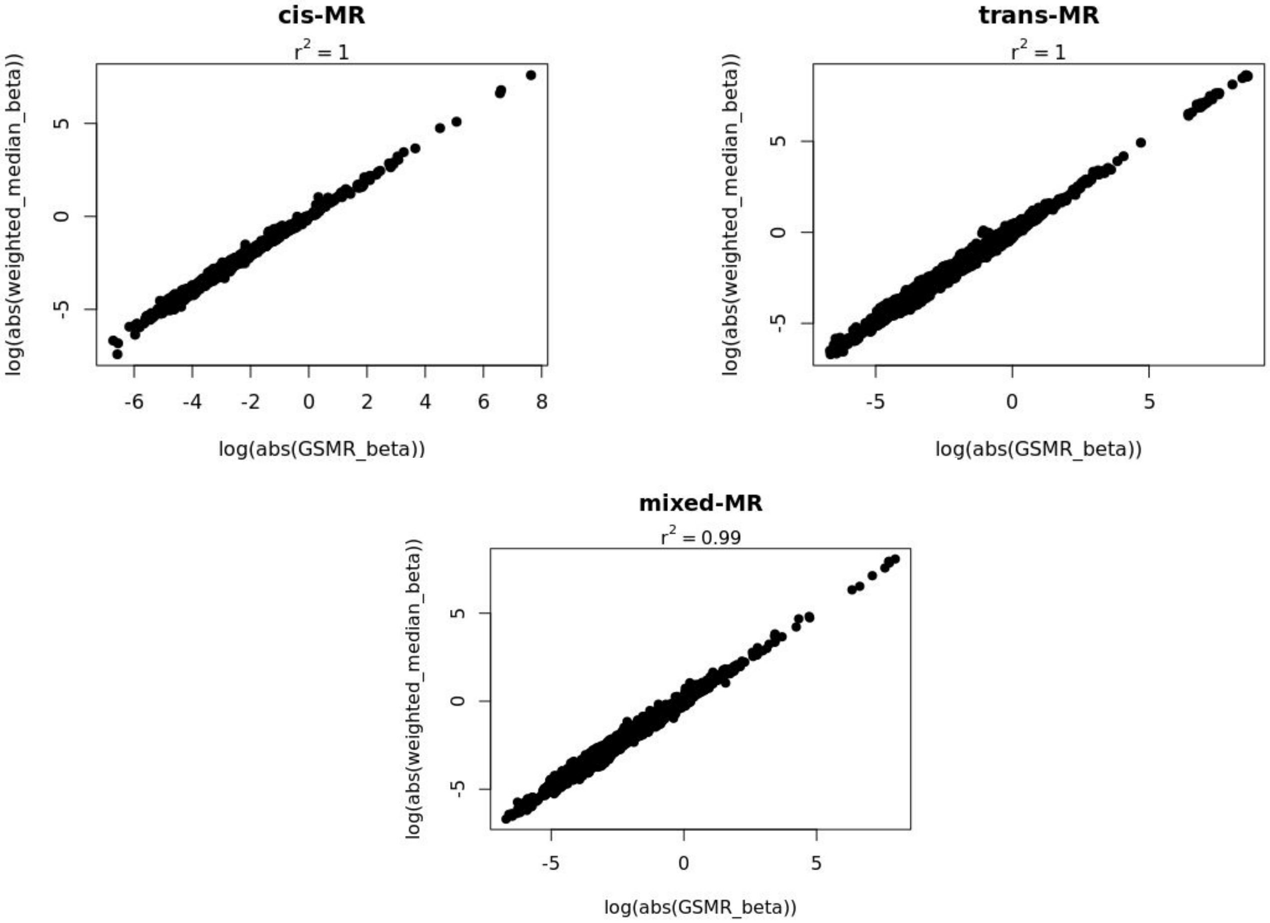

**Supplementary Figure 2 : Use of Protein-protein interactions to expand the spectrum of druggable targets**

### Approach

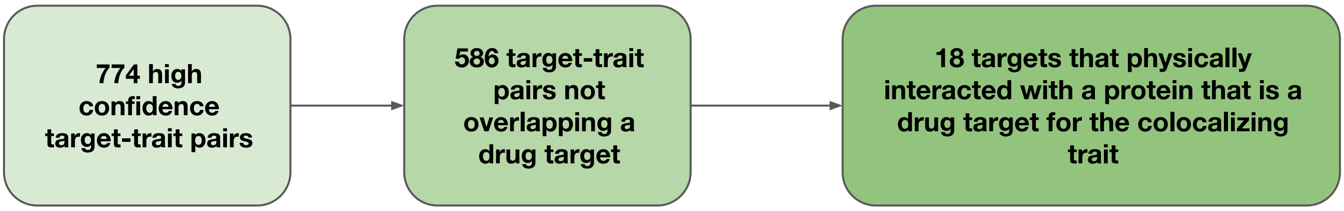


### Selected examples

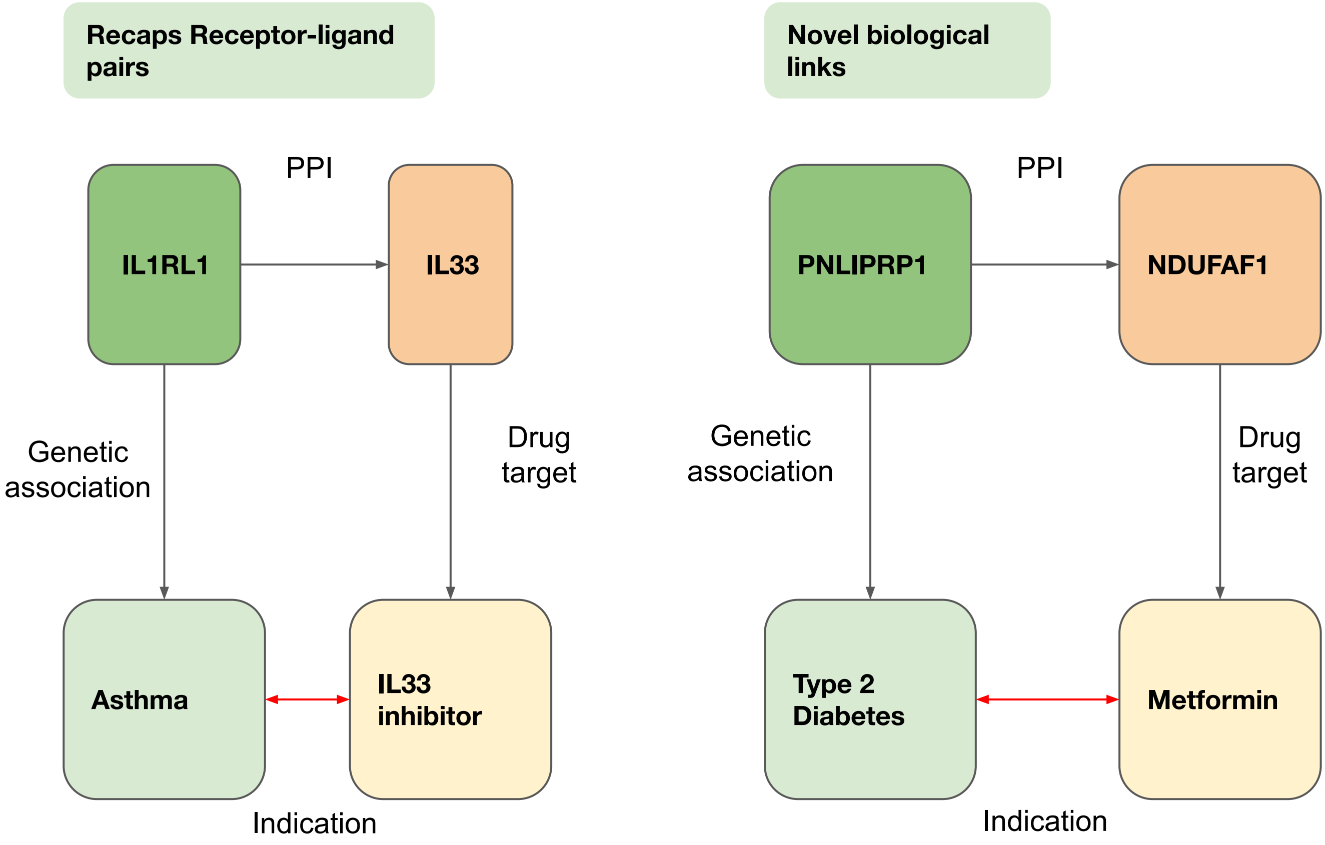

**Supplementary Figure 3: Trans-colocalization between IBD and ALPI at *FUT2***

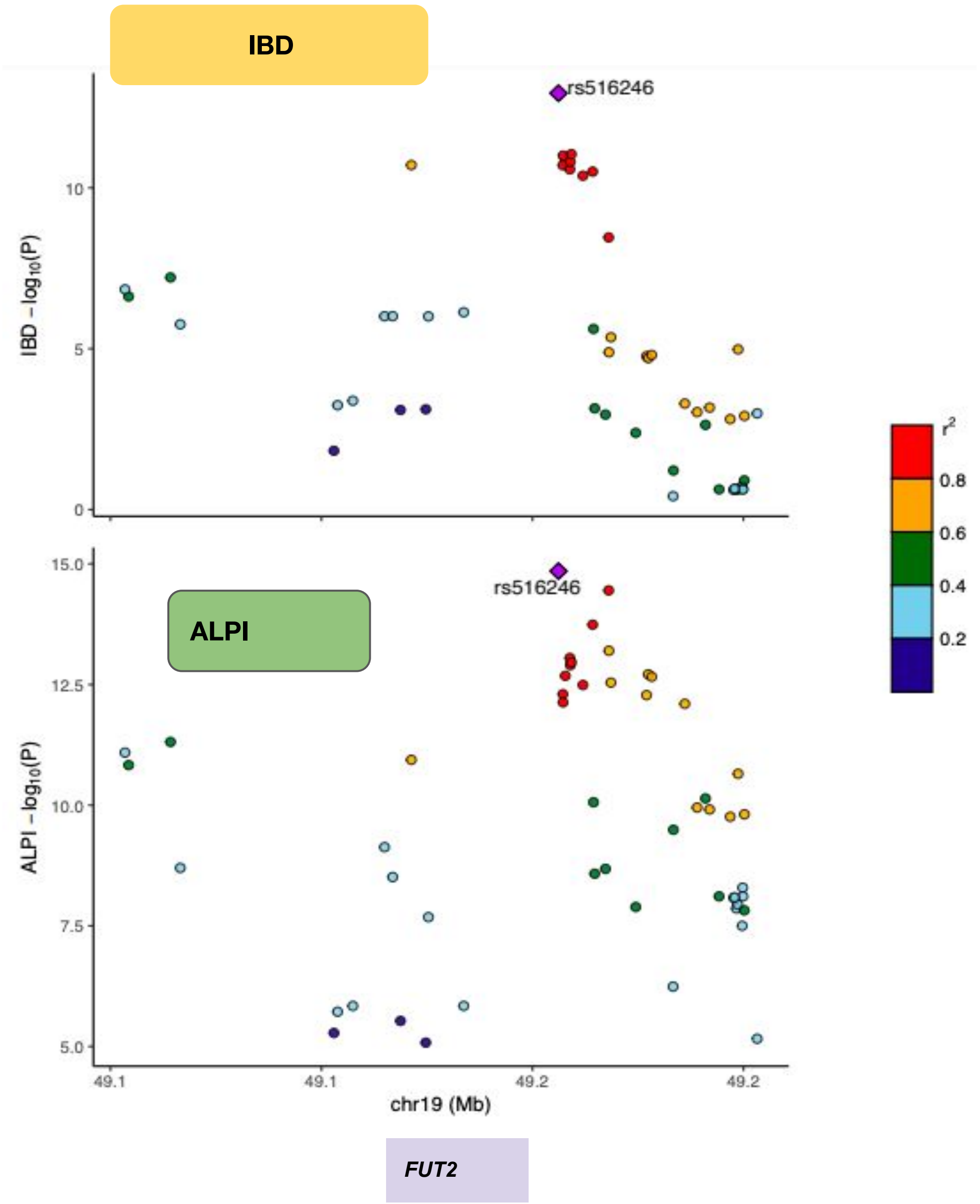

**Supplementary Figure 4: Forest plot showing the directionally opposing associations of asthma and IBD with different genetically predicted higher plasma proteins**

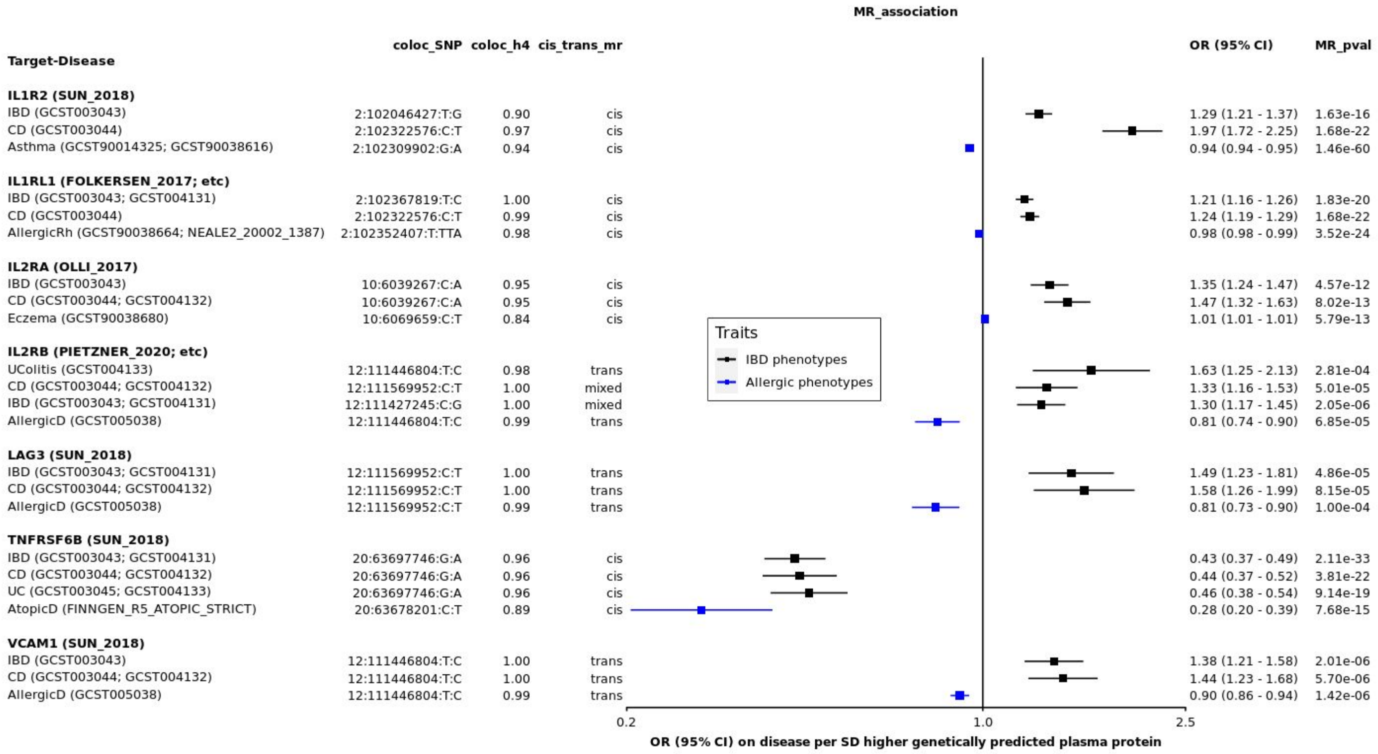

**Supplementary Figure 5: Difference between effect estimates of cis- and trans-pQTLs**

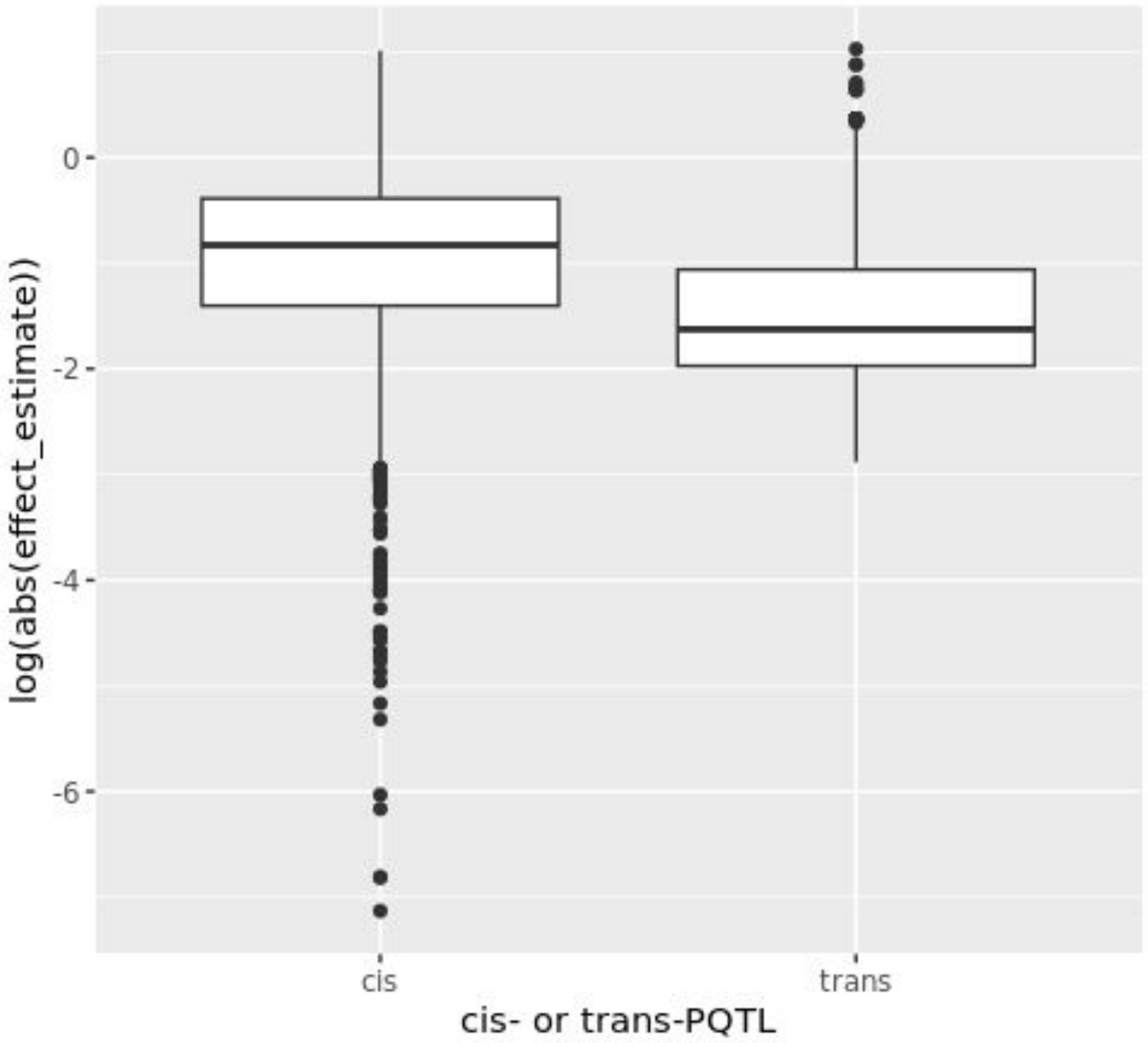
